# Physics of Virus Transmission by Speaking Droplets

**DOI:** 10.1101/2020.05.12.20099630

**Authors:** Roland R. Netz, Adriaan Bax, William A. Eaton

## Abstract

Droplets of oral fluid emitted by speaking are a long-recognized mechanism of respiratory virus transmission. While there have been many simulations of droplet evaporation to determine whether droplets containing virions remain floating in air or rapidly fall to the ground, they typically conceal the fundamental mechanisms because of the use of numerics. To make the physics of emitted oral fluid droplets easily understood, we present simple and transparent algebraic equations that capture the essential physics of the problem. Calculations with these equations provide a straightforward way to determine the airborne lifetime of emitted droplets after accounting for the decrease in droplet size from water evaporation. At a relative humidity of 50%, droplets with initial radii larger than about 50 μm rapidly fall to the ground while smaller, potentially-virus containing droplets shrink in size and remain airborne for many minutes. Rough estimates of airborne virion emission rates while speaking support the proposal that covering the mouth can help end the pandemic more quickly.

---

The physics of water droplets is a well-studied subject and its relevance to virus and transmission long known (*1-12*). It is a subject that has aroused renewed interest because of the Covid-19 pandemic and has motivated scientists to perform new kinds of experiments. Just published laser light-scattering experiments of Anfinrud *et al*. (*13*) show that the number of oral fluid droplets are emitted into the air while speaking is orders of magnitude larger than previously detected using less sensitive methods (9) and that blocking such droplets is easily accomplished with a cloth mouth cover (13)(youtu.be/qzARpgx8cvE). Previous physics calculations of droplet evaporation to determine whether droplets containing viruses remain floating in air or rapidly fall to the ground typically involve numerical simulations, which hide the fundamental mechanisms (*5,14,15*). In addition, the mathematics employed is too complex to be understood by other than physical scientists. We have investigated all aspects of this physics problem and present simple and transparent algebraic equations that capture the essential physics.

Our equations answer two important questions. First, how long does it take for a virus-containing droplet of a given size to fall to the ground by gravity to potentially contaminate a surface? Second, for each relative humidity, how much time does it take for water evaporation to reduce a virus-containing droplet to a size that leaves it floating in air for a sufficiently long time to allow direct transmission of the virus to another person? The answer to the first question is easily obtained by simply equating the gravitational and Stokesian viscous forces on a falling object (*mg* = *6πηRv*) to obtain the terminal velocity (*v*). This simplistic treatment is justified in ref. (*16*). The mean time for a particle to reach the ground is then:

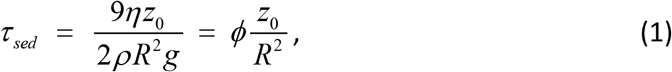

where *τ_sed_* is the mean time for a droplet of radius, *R*, to reach the ground from a height, z_0_, with both *R* and z_0_ in units of μm (1 μm = 10^−6^ m). The prefactor, *ϕ* = *9η/*(*2ρg*) *=* 0.85 × 10^−2^ μm s, is calculated from the viscosity of air at 25°C, *η* = 1.86 × 10^−8^ g/μm s, water density, *ρ* = 10^−12^ g/μm^3^, and the gravitational constant, *g* = 9.8 × 10^6^ μ.m /s^2^. A few examples are instructive. Droplets placed initially at 1.5 m (the average height above ground for the mouth of a standing human adult) with radii of 1 μm, 10 μm, or 100 μm will require 1.3×10^4^ s (~ 3.5 hours), 130 s and 1.3 s, to settle to the ground, respectively.

Determining whether or not a virus-containing droplet will remain airborne to cause an infection requires determination of the rate of evaporation of water, which is a more complex problem and is different for different size regimes. The most important effects to consider in the size regime of interest are the cooling of the droplet from the heat loss due to water evaporation, which slows the evaporation rate, and the osmotic effect of the soluble droplet contents, that reduces the water vapor pressure at the droplet surface and therefore decreases the evaporation rate. Oral fluid, which is primarily saliva, contains about 99% by volume water, with the remaining 1% of volume consisting of proteins, mucus, epithelial cells, white cells, electrolytes, small molecules (*17*), and possibly a virion. We assume that ~1% volume of water is strongly bound to the solutes. The volume fraction of non-volatile solute, which includes the strongly bound hydration water, is then about Φ_0_ = 0.02. Dehydration of the droplet in less than water vapor-saturated air results in a so-called “droplet nucleus” (*dn*), which retains additional water. Balance of the water vapor pressure of the droplet and the ambient air results in an equilibrium solute volume fraction of Φ*_dn_* = 1 - RH, where *RH* is the relative humidity. Thus, for *RH* = 0.5, the initial droplet radius can maximally decrease due to evaporation by about a factor of ((1 - *RH*)/ Φ_0_)^1/3^ ≈ 3.

The 3 different size regimes that require different theoretical treatments are: droplet radii *R* < 100 nm, 100 nm < *R* < 60 μm and > 60 μm. We shall only be concerned with the latter 2 regimes, as < 100 nm radii are in the size regime of a single virion (*τ_sed_* = several days), which cannot be emitted without a surrounding layer of oral fluid. In the following, we assume that the droplet has escaped from any surrounding water vapor cloud (*8*) to be in ambient air. In the intermediate size regime of 100 nm < *R* < 60, the result for *τ_ev_*, the time it takes for complete evaporation of a pure water droplet of initial radius R_0_, including cooling, is (*16*)

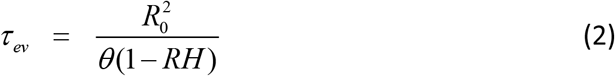

where *RH* is the relative humidity, *θ* = *2αD_w_c_g_v_w_* = 3.7 × 10^2^ μm^2^/s at 25^0^C is a constant with units of diffusion and the numerical prefactor, *α =* 0.32, accounts for evaporation-cooling effects (*16*). The diffusion constant for a water molecule in air, *D_w_*, is 2.82 × 10^7^ μm^2^/s, the water number concentration, *c_g_*, in saturated air is 6.6 × 10^5^ μm^−3^, and the water molecular volume, *v_w_*, in liquid water is 3.07 × 10^−11^ μm^3^, all at 25°C. Notice that the quadratic dependence of *τ_ev_* on R_0_, indicates that evaporation accelerates as the droplet gets smaller and therefore falls more slowly.

The theory is more complex for inclusion of the osmotic effect of the contents of a droplet, the droplet nucleus. In this case, the mean time for a droplet of initial radius R_0_ to shrink to a radius *R* from water evaporation, is given by (*16*)

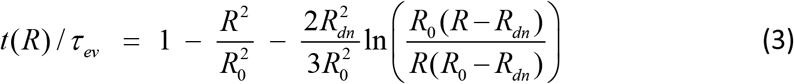

where *R_dn_* is the equilibrium droplet nucleus radius, which we estimated above to be ~R_0_/3. The last term in equation (3) accounts for the vapor pressure reduction due to solutes. At *R* < 1.5 *R_dn_*, the evaporation time enters the solute-dominated regime and diverges, albeit only logarithmically, in the limit *R* → *R_dn_* (*16*). Therefore, for times prior to achieving perfect equilibrium the logarithmic term is small enough to be neglected and equation 3 simplifies to

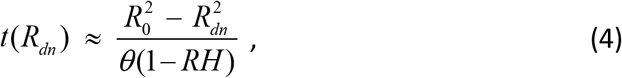

where. At a relative humidity of *RH* = 0.5, a common value for room air, the mean evaporation times for droplets with initial radii, R_0_, of 1, 10, and 100 μm are 5 ms, 0.5 s, and 50 s, while the corresponding sedimentation mean times, *τ_sed_*, from equation (1) are 1.3 × 10^4^ s, 130 s, and 1.3 s. Consequently, the 1 and 10 μm droplets will dry out and stay floating for even longer, which will be determined by the radius of the droplet nuclei, *R_dn_*. Thus, droplets with an initial radius of *R*_o_= 20 μm will shrink to a droplet nucleus radius of ~7 μm in *t*(*R_dn_*) ≈ 2 s (equation 4), with the droplet nuclei remaining airborne for about 4 minutes (equation 1).

It is useful to define a “critical radius”, 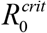, where the evaporation and settling times are equal, i.e. 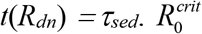 is obtained by combining equations (1) and (3) (with *R_dn_* = *R*_0_/3) to give:

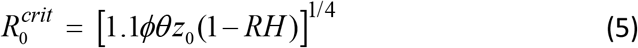

For *RH* = 0.5 and z_0_ = 1.5 m, the critical radius is 40 μm. This means that droplets with radii greater than 40 μm will fall to the ground before drying out, while droplets with radii less than 40 μm will remain floating in the air. A more accurate value for the critical radius of ~50 μm is obtained by solving equations that take into account evaporation of droplets *while* sedimenting (*16*).

Equation (3) does not apply to the third size regime of *R*_o_ > 60 μm. At *R*_o_ > 60 μm, calculating the evaporation rate for droplets at rest (the so-called stagnant approximation) is no longer valid and the evaporation is accelerated by the air flow around the droplet. This effect can be treated by assuming a concentration boundary layer around the droplet. At the same time, non-linear hydrodynamic effects produce a flow boundary layer around the droplet outside of which viscosity effects are negligible. Using double boundary layer theory (*16*), the evaporation for *R*_o_ > 60 μm is shown to be even faster than predicted by equation (3). Since these modifications only become important for larger radii than our equation (3) predicts, the estimate of the critical droplet size of 40 μm is not modified by these complex hydrodynamic effects.

Can we say anything useful about the number of emitted virions while speaking? Anfinrud *et al*. used their sensitive laser light scattering method to count droplets in each 1/60^th^ s frame of their video, which yields an approximate average droplet emission rate of ~10^3^/s (*13*). We also need to know the concentration of virions in saliva and the volume of the droplets. Wölfel *et al*. reported an average virion concentration of 7×10^6^ (*18*) virions/cc. There are no reported values for the average volume at droplet count rates comparable to the findings of Anfinrud *et al*. However, all of the distributions determined since the work of Duguid (*2*) show that the vast majority of droplet diameters are less than 50 microns (*9*) (and references therein). We can therefore get an informative estimate of the possible range for the rate of emitted airborne virions by calculating this number for each assumed initial droplet radius.

Given the many contents of saliva, in addition to the possibility of a virion, we estimate that the smallest possible radius before evaporation is ~1 μm. Table 1 shows the calculated values for initial droplet radii (*R*_0_) from 1 to 20 μm, which predicts that the number of emitted virions while continuously speaking ranges from 2 to ~10^4^ per minute. Comparing the evaporation times at a relative humidity of 50% with the sedimentation times in Table 1 shows that for all radii in this range droplet nuclei remain airborne for times sufficiently long that their airborne lifetime will be determined by the turnover time of the air handling system. It is not known what fraction of the virions in these concentration measurements are infectious, but it has been argued that in some systems as few as a single active virion can cause an infection (*19*). The very large range of virion emission rates in Table 1 calls for both an accurate determination of the fraction of airborne virions that are infectious, as well as accurate droplet size distributions at the high rate of emission determined by laser light scattering (*13*). Measurements of droplet sizes at these high rates are currently underway at NIH.

**Table 1.** Theoretical virion emission rates (*k*), sedimentation times (*τ_sed_*) at z_0_ = 1.5 m, and evaporation times (*t*(*R_dn_*)) at 25^0^C for initial radii (R_0_) and for droplet nuclei at 50% relative humidity (*RH* = 0.5).

| $R_0$<br>($\mu\text{m}$) | $k^1$<br>(virions/min) | $t(R_{dn})^2$<br>(min) | $\tau_{sed}$ (droplet, $R_0$)<sup>3</sup><br>(min) | $\tau_{sed}$ (droplet nuclei, $R_0/3$)<sup>3</sup><br>(min) |
| --- | --- | --- | --- | --- |
| 1 | 2 | $8 \times 10^{-5}$ | 200 | $2 \times 10^3$ |
| 3 | 50 | $8 \times 10^{-4}$ | 20 | 200 |
| 5 | 200 | $2 \times 10^{-3}$ | 9 | 80 |
| 10 | $2 \times 10^3$ | $8 \times 10^{-3}$ | 2 | 20 |
| 20 | $1 \times 10^4$ | $3 \times 10^{-2}$ | 0.5 | 5 |
| 40 | $1 \times 10^5$ | 0.1 | 0.1 | - |
| 80 | $9 \times 10^5$ | 0.5 | 0.03 | - |

Overall, the above analysis strongly supports the concept that simply speaking can be a major mechanism of Covid-19 transmission and that covering the mouth in public, as suggested by the work of Anfinrud *et al* (*13*) (youtu.be/qzARpgx8cvE) and others (*12, 20*), could help to more rapidly contain and potentially end the pandemic. It also suggests that mouth covering is a more effective way of reducing transmission than maintaining a distance of at least 6 feet from another person. This conclusion is strongly supported by epidemiological evidence. The majority of the 24 million Taiwanese have been wearing masks since the first infections were discovered, which almost certainly has contributed to the low number of deaths – only 6 reported in Taiwan as of April 30 (with only 1 in the past 2 weeks) compared to more than 25,000 deaths in the state of New York with 4 million fewer inhabitants. Even more convincing is Vietnam, where the total number of reported deaths is zero and wearing a mask is mandatory in public places for the entire population of 94 million. Finally, Austria and Germany, where face masks are required, are the only large non-Asiatic countries where there is marked slowing in the rate of new cases (https://coronavirus.ihu.edu/map.html).

## Data Availability

All data contained in uploaded manuscript

## Acknowledgement

This work was supported by the European research council (ERC) via an Advanced Investigator Grant and the intramural research program of the National Institutes of Diabetes and Digestive and Kidney Diseases of the National Institutes of Health.

## Notes

### Competing Interest Statement

The authors have declared no competing interest.

